# Comparison of Rupture Risk Between Saccular and Fusiform Abdominal Aortic Aneurysms Using a National Clinical Database in Japan

**DOI:** 10.1101/2023.06.06.23291061

**Authors:** Hirotsugu Ozawa, Arata Takahashi, Ryuzo Bessho, Katsuyuki Hoshina, Kota Shukuzawa, Takao Ohki

## Abstract

**Background:** Saccular AAAs are thought to pose an elevated risk of rupture, but not much is known about the extent of this risk. Therefore, we aimed to conduct a survey of saccular abdominal aortic aneurysms (AAAs) and to compare the risk of rupture between fusiform and saccular AAAs.

**Methods:** We performed a retrospective cohort study on patients who underwent primary endovascular repair for a degenerative AAA between 2016 and 2019, and who were registered in the National Clinical Database in Japan.

**Results:** A total of 27,290 patients were included in the study. Of these, 7.8 % (n=2142) had saccular AAAs and the remaining 92.2% (n = 25,148) were fusiform. In addition, 4.3% (n = 92) of saccular AAAs and 5.4% (n = 1351) of fusiform AAAs were ruptured. Saccular AAAs ruptured at smaller dimeters than fusiform AAAs (median, 55.6 mm vs 68.0 mm, p < 0.001), and were operated on at smaller diameters than fusiform AAAs in non-ruptured cases (median, 44.0 mm vs 51.0 mm, p < 0.001). The rupture rate was significantly higher in saccular AAAs than in fusiform AAAs in the 40-54 mm diameter range, in which saccular morphology was found to be an independent risk factor for rupture by adjusting for gender and aneurysm diameter (odds ratio, 2.54, 95% confidence interval, 1.75-3.69). In addition, receiver-operating characteristic analysis revealed that the cut-off diameter to predict rupture was smaller in saccular AAAs than in fusiform AAAs (50.5 mm and 59.5 mm, respectively).

**Conclusion:** Saccular AAAs are more prone to rupture than fusiform AAAs in the 40-54 mm diameter range, which supports the idea that saccular AAAs should be treated at smaller diameters. The 9.0 mm difference in the predicted diameters for the rupture between fusiform and saccular AAAs suggests that the threshold diameter for intervention of saccular AAAs can be set approximately 1 cm smaller than that of fusiform AAAs.

**Clinical Perspective:** *What Is New?:*

- Saccular abdominal aortic aneurysms (AAAs) ruptured at smaller dimeters than fusiform AAAs (median, 55.6 mm vs 68.0 mm, p < 0.001), and receiver-operating characteristic analysis revealed that the cut-off diameter to predict rupture was smaller in saccular AAAs than in fusiform AAAs (50.5 mm and 59.5 mm, respectively).
- In AAAs with a dimeter of 40-54 mm, saccular morphology turned out to be an independent risk factor for rupture by adjusting for gender and aneurysm diameter (odds ratio, 2.54, 95% confidence interval, 1.75-3.69).

*What Are the Clinical Implications?:*

- Saccular AAAs are more prone to rupture than fusiform AAAs in the 40-54 mm diameter range, which supports the current idea that saccular AAAs should be treated at smaller diameters.
- The 9.0 mm difference in the predicted diameters for the rupture between fusiform and saccular AAAs suggests that the threshold diameter for intervention of saccular AAAs can be set approximately 1 cm smaller than that of fusiform AAAs.

## INTRODUCTION

Saccular abdominal aortic aneurysms (AAAs), defined as asymmetric enlargement of the aorta, account for only about 5% of all AAAs, with the majority being fusiform AAAs.^1–3^ It has long been believed that saccular aneurysms are more prone to rupture.^2, 4–7^ However, to date, not much is known about the natural history and the risk of rupture in saccular AAAs.

Current international guidelines recommend elective repair for AAAs with a diameter ≥ 55 mm in men and ≥ 50 mm in women, but this statement is limited to fusiform AAAs.^8, 9^ For saccular AAAs, however, these guidelines suggest elective repair at a smaller diameter, but fail to provide a size threshold for intervention. Thus, the optimal management of saccular AAAs is unclear, and surgeons assess the risk of rupture and determine the indications for elective repair on a case-by-case basis.

Cohort studies reporting on the clinical management of saccular AAA has been limited.^10, 11^ According to a recent cohort study of saccular AAAs from the Netherlands,^11^ saccular AAAs were operated on at smaller diameters in the elective setting and became symptomatic/ruptured at smaller diameters than fusiform AAAs. The authors also added that a diameter of 45 mm seems to be an acceptable threshold. However, the number of symptomatic/ruptured cases of saccular AAAs with a dimeter < 45 mm in this study was insufficient for a powerful statistical analysis.

In the current study, we conducted a survey of saccular AAAs that were treated with endovascular aortic repair (EVAR) in Japan, and we sought to compare the risk of rupture between fusiform and saccular AAAs, using data registered in the National Clinical Database (NCD) in Japan.

## METHODS

This study was approved by the Institutional Review Board at The Jikei University School of Medicine (33-189[10806]). Informed consent was waived for this study. The study protocol was registered with the University Hospital Medical Information Network Clinical Trials Registry (UMIN-CTR; UMIN000050383).

### Database

The NCD in Japan, which was launched in 2010 and commenced patient registration in 2011, is a nationwide prospective registry that can collect data on surgical procedures from more than 5000 institutions throughout Japan and has very high coverage because of its link with the surgeon/hospital certification system.^12^ In addition, previous studies have verified the data quality of the NCD.^13–15^ Regarding EVAR procedures for AAAs, the Japanese Committee for Stentgraft Management (JACSM), established in December 2006 to ensure safe and appropriate use of commercial stent grafts, has started a nationwide EVAR registry from 2007, using a web-based case-registry form.^16^ Participating institutions were obligated to register detailed data, including preoperative findings on AAAs, operative findings and postoperative outcomes of EVAR. Since 2016, through the collaboration between the JACSM and the NCD, the data registration is now done on the NCD website.

### Inclusion and Exclusion Criteria for Data

Patients undergoing primary EVAR for a degenerative AAA in Japan between January 2016 and December 2019 were included in the study. Cases of AAA with concomitant iliac artery aneurysm, dissecting/inflammatory/mycotic AAA, or AAA with vasculitis/connective tissue disease were not included in the study. Additionally, cases were not included if the AAA was treated with snorkel/chimney, fenestrated/branched, or debranching EVAR. Patients with AAA who underwent open surgical repair (OSR) were not included because it was not required to register AAA morphology in the NCD for such cases. Those with an aneurysm diameter < 25 mm were also excluded because the suggested reporting standard in the guidelines states that the definition of AAA, which is ≥ 30 mm in diameter in men, should be lower in women and in the Asian population and therefore suggests an exceptional situation.^9^

### Collected Data

Data registered into the NCD for each patient included age, sex, comorbidities, and the etiology, anatomical factors and clinical status of the AAA. Comorbidities registered included hypertension, diabetes mellitus, coronary artery disease, cerebrovascular disease, renal dysfunction (estimated glomerular filtration rate < 60 ml/min/1.73m^2^), and respiratory disorder. Anatomical factors included the shape of the AAA (fusiform or saccular), and aneurysm diameter; maximum minor-axis diameter chosen if fusiform and maximum transaortic diameter if saccular. Status of the AAA was described according to the existence of rupture (non-ruptured or ruptured). These data were confirmed by each surgeon.

### Outcomes

The primary outcome was the aneurysm diameter at which saccular AAAs were operated on by EVAR in the non-ruptured and ruptured cases. The secondary outcomes included the odds ratio (OR) of saccular morphology for becoming ruptured and a cut-off value of aneurysm diameter for predicting the rupture of a saccular AAA.

### Statistical Analyses

Patients were stratified according to the shape of the AAA (fusiform versus saccular), and according to the clinical status (non-ruptured versus ruptured). We obtained data on the aneurysm diameter at which saccular AAAs were operated on by EVAR in the non-ruptured and ruptured cases. Then, we compared the rupture rate by aneurysm diameters of fusiform and saccular AAAs utilizing categorical variables for each 5 mm diameter. The rupture rate in this study was the likelihood of repair for rupture, defined as the number of ruptured cases over the total number of cases. The OR for rupture was determined by adjusting for all variables included in the guidelines as indications for repair, namely, sex, aneurysm diameter, and aneurysm morphology, except for growth rate that was not captured in the NCD. Furthermore, receiver-operating characteristic (ROC) analysis was performed to evaluate the predicting power of rupture for saccular AAAs to rupture as well as that for fusiform AAAs.

Categorical variables are presented as numbers and percentages, and continuous variables are presented as mean and standard deviations or median and interquartile ranges (IQR). Categorical variables were compared with a chi-square test, and continuous variables were compared using a *t* test or Mann-Whitney *U* test when appropriate. To compare the risk of rupture between fusiform and saccular AAAs, an OR was determined per diameter category using logistic regression analysis. All statistical analyses were performed using SPSS Statistics version 27 (IBM, Armonk, NY), and *p* < 0.05 was considered statistically significant.

## RESULTS

From January 2016 to December 2019, all 27,418 patients who underwent primary standard EVAR for degenerative AAAs were registered in the NCD. Those with AAA diameter < 25 mm (n = 128) were excluded from the study. Finally, a total of 27,290 patients were included in the study.

### Patient Characteristics

Patient characteristics including the morphology and the clinical status of AAA at treatment are shown in Table 1. Among the 27,290 cases that were included in this study, 7.8% (n = 2142) of AAAs were saccular and the remaining 92.2% (n = 25,148) were fusiform, while 5.3% (n = 1443) were ruptured and the remaining 94.7% (n = 25,847) were non-ruptured. Specifically, 4.3% (n = 92) of saccular AAAs and 5.4% (n = 1351) of fusiform AAAs were ruptured. At operation for non-ruptured case, the aneurysm diameter was significantly smaller in saccular AAAs than in fusiform AAAs (median, 44.0 mm vs 51.0 mm, *p*< 0.001). Similarly, aneurysm diameter at rupture was significantly smaller in saccular AAAs than in fusiform AAAs (median, 55.6 mm vs 68.0 mm, *p* < 0.001). Comparing the non-ruptured and ruptured cases, sex, coronary artery disease, renal dysfunction, and respiratory disorder were significantly different in the fusiform AAAs, while renal dysfunction and respiratory disorder were significantly different in the saccular AAAs. Ratio of women tended to be higher in ruptured cases than in non-ruptured cases in fusiform AAAs, but not in saccular AAAs.

**Table 1.**
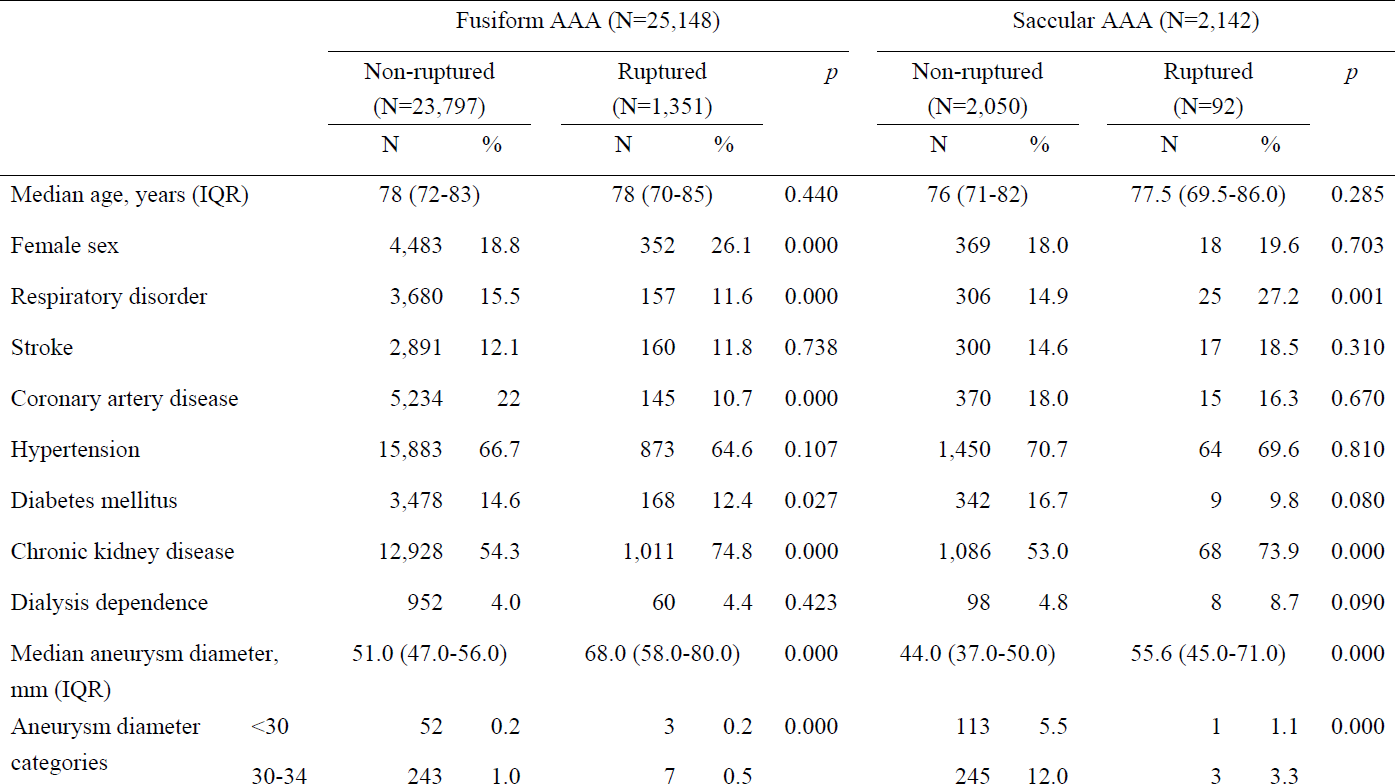
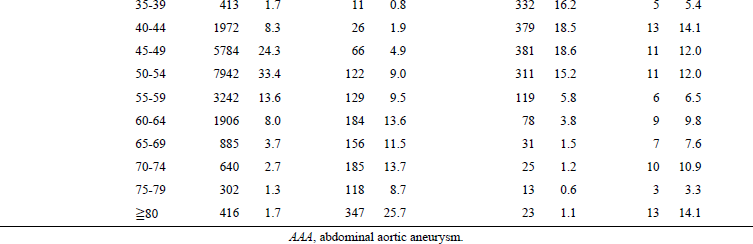
Patient characteristics and data of abdominal aortic aneurysms.

### Comparison of the Rupture Risk Between Fusiform and Saccular AAAs

In Table 2, comparison of the rupture rate was made by aneurysm diameter between fusiform and saccular AAAs using categories of 5-mm diameter increments. In AAAs with aneurysm diameter of 40-54 mmm, each category was significantly more likely to rupture in saccular AAAs than in fusiform AAAs. On the other hand, in AAAs with aneurysm diameters of 30-39 mm and 55-69 mm, there was no statistically significant difference in the rupture rates between fusiform and saccular AAAs.

**Table 2.** Comparison of rupture rate between fusiform and saccular abdominal aortic aneurysms.

| Aneurysm diameter | Fusiform AAA |  |  | Saccular AAA |  |  | OR | 95% CI |  | *p* |
| --- | --- | --- | --- | --- | --- | --- | --- | --- | --- | --- |
|  | Total | Rupture | % | Total | Rupture | % |  | Lower | Upper |  |
| 30-34 | 247 | 4 | 1.6 | 248 | 3 | 1.2 | 0.744 | 0.165 | 3.359 | 0.700 |
| 35-39 | 424 | 11 | 2.6 | 337 | 5 | 1.5 | 0.565 | 0.195 | 1.643 | 0.295 |
| 40-44 | 1998 | 26 | 1.3 | 392 | 13 | 3.3 | 2.602 | 1.325 | 5.108 | 0.005 |
| 45-49 | 5850 | 66 | 1.1 | 392 | 11 | 2.8 | 2.530 | 1.325 | 4.831 | 0.005 |
| 50-54 | 8064 | 122 | 1.5 | 322 | 11 | 3.4 | 2.303 | 1.230 | 4.312 | 0.009 |
| 55-59 | 3371 | 129 | 3.8 | 125 | 6 | 4.8 | 1.267 | 0.548 | 2.931 | 0.580 |
| 60-64 | 2090 | 184 | 8.8 | 87 | 9 | 10.3 | 1.195 | 0.590 | 2.422 | 0.621 |
| 65-69 | 1041 | 156 | 15.0 | 38 | 7 | 18.4 | 1.281 | 0.554 | 2.960 | 0.562 |

AAA, abdominal aortic aneurysm; OR, odds ratio; CI, confidence interval.

In addition, focusing on ruptured cases, Figure 1 shows the distribution of ruptured cases by diameter category in fusiform and saccular AAAs, suggesting that saccular AAAs may rupture at smaller diameters than fusiform AAAs.

**Figure 1.**
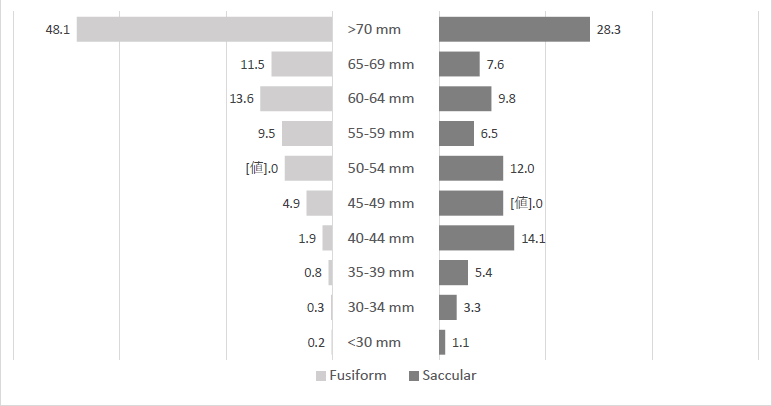
Distribution of ruptured aneurysms by diameter category in fusiform and saccular abdominal aortic aneurysms (AAAs). The numbers next to the bars represent the percentages of rupture cases in each diameter category among all rupture cases of fusiform or saccular AAAs.

As shown in Table 3, risk analysis of rupture was performed for small (30-39 mm in diameter), medium (40-54 mm in diameter), and large (55-69 mm in diameter) AAAs, adjusted for sex, aneurysm shape, and diameter category. As a result, saccular shape turned out to be an independent risk factor for rupture in medium AAAs (OR, 2.54, 95% confidence interval [CI], 1.75-3.69), but not in small and large AAAs (OR 0.62, 95% CI, 0.26-1.47 and OR 1.28, 95% CI, 0.81-2.02, respectively). In addition, female sex was identified as an independent risk factor for rupture in all AAAs except small AAAs, and diameter category was identified as an independent risk factor for rupture only in large AAAs.

**Table 3.**
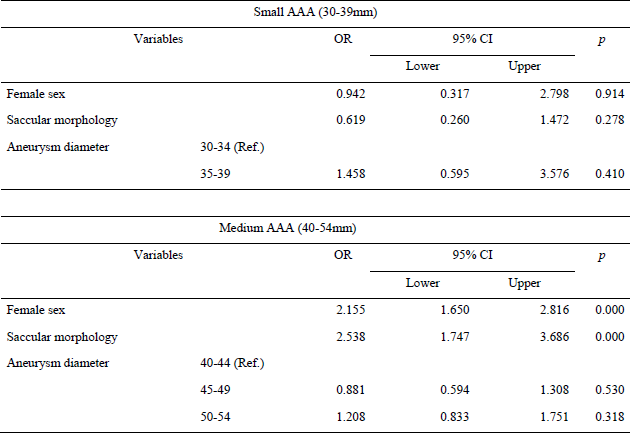
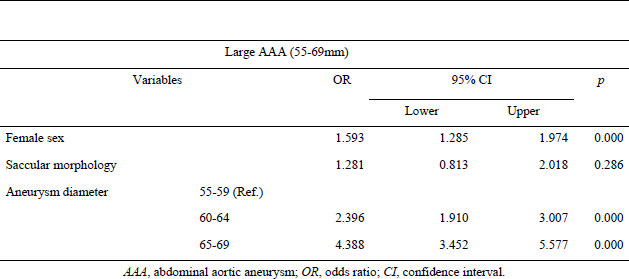
Adjusted odds ratio for rupture in small, medium, and large abdominal aortic aneurysms.

### ROC Analysis to Predict Rupture of Fusiform and Saccular AAAs

Diameters that predict rupture in fusiform and saccular AAAs were analyzed using ROC analysis and are shown in Figure 2 and Table 4. The area under the curve of the diameter that predicts rupture of fusiform and saccular AAAs was 0.830 and 0.752, respectively. A cut-off diameter with the highest predictive power for rupture was 59.5 mm in fusiform AAAs (sensitivity 73.4%, specificity 82.5%) and 50.5 mm in saccular AAAs (sensitivity 63.0%, specificity 77.7%). If the cut-off diameter was set at 55 mm in fusiform AAAs, the sensitivity and specificity for predicting rupture were 79.8% and 73.7%, respectively. As for saccular AAAs, if the cut-off diameter was set at 45 mm, the sensitivity and specificity were 71.7% and 58.1%, respectively. Furthermore, the sensitivity of the cut-off diameter of 55 mm in fusiform AAAs (79.8%) was comparable with that of 43 mm in saccular AAAs (78.3%).

**Figure 2.**
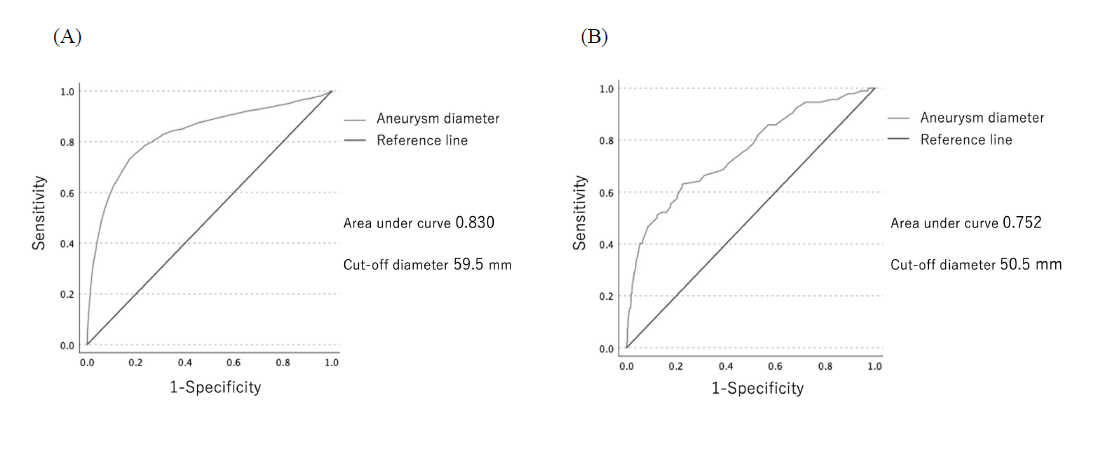
Receiver-operating characteristic analysis of rupture for (A) fusiform and (B) saccular abdominal aortic aneurysms (AAAs). The area under the curve of the diameter that predicts rupture of fusiform and saccular AAAs was 0.830 and 0.752, respectively. A cut-off diameter with the highest predictive power for rupture was 59.5 mm in fusiform AAAs and 50.5 mm in saccular AAAs.

## DISCUSSION

In this retrospective cohort study, among patients who underwent primary EVAR for degenerative AAA between 2016 and 2019, 7.8% had a saccular AAA. Saccular AAAs ruptured at smaller dimeter than fusiform AAAs and were operated on at smaller diameters than fusiform AAAs in non-ruptured cases, which supports the current treatment guidelines for AAAs.^8, 9^ The rupture rate was significantly higher in saccular AAAs than in fusiform AAAs in the medium-size category, in which saccular morphology turned out to be an independent risk factor for rupture by adjusting for gender and aneurysm diameter. In addition, the cut-off diameter for predicting rupture was 9.0 mm smaller in saccular AAAs than in fusiform AAAs in the ROC analysis.

The majority of AAAs are fusiform AAAs and often occur as a result of degeneration of the aortic wall. On the other hand, saccular AAAs are rare and seem to be mainly caused by degeneration, followed by a variety of etiologies such as dissection, trauma, infection, and vasculitis.^10^ Whereas aneurysm diameter and growth rate have been widely accepted as major indications for repair of an AAA, a saccular morphology has also been considered as an indication for repair. Despite the common perception that saccular aneurysms are at high risk of rupture, not much has been known about the natural history and the risk of rupture in saccular AAAs. Furthermore, there are limited data on what diameter saccular AAAs are treated with surgery in clinical practice.

According to a previous large cohort study of saccular AAAs conducted in the Netherlands,^11^ saccular AAAs were operated on at smaller diameters than fusiform AAAs in the elective setting (mean, 53.0 mm vs 61.0 mm, *p* < 0.001) and became symptomatic and/or ruptured at smaller diameters than fusiform AAAs (mean, 70.7 mm vs 76.5 mm, *p* = 0.033). The authors also added that a diameter of 45 mm seems to be an acceptable threshold for surgery, based on the finding that the proportion of symptomatic/ruptured patients was similar between saccular AAAs with diameters < 45 mm and fusiform AAAs with diameters < 55 mm. However, the number of symptomatic/ruptured cases of saccular AAAs (n = 83), especially those with a diameter < 45 mm (n = 7), was insufficient for a powerful statistical analysis. In the present study, we decided to focus on the rupture of AAAs, since patients could be described as symptomatic if the aneurysm caused a pulsing sensation or local compression symptoms, and such symptomatic patients should be differentiated from patients presenting with abdominal or back pain. Furthermore, the primary goal of the physician taking care of AAA patient is to predict the risk of rupture, not the development of symptoms.

In the present study, the percentage of saccular AAAs out of all treated AAAs was 7.8%, which is similar to previous reports (approximately 5%). The median aneurysm diameter at rupture was smaller in saccular AAAs than fusiform AAAs (fusiform AAAs: 68.0 mm; saccular AAAs: 55.6 mm), suggesting that saccular AAAs are more prone to rupture.

The median diameters at rupture in our study were smaller than those at symptom/rupture in the previous study form the Netherlands (fusiform AAAs: 75.0 mm; saccular AAAs: 68.0 mm). However, this may be due to the smaller aortic diameter in the Asian populations,^17^ and also due to the predisposition to symptomatic/ruptured presentation at smaller diameters in the Asian population.^18^

The present study suggests that if AAAs are classified by size, each size range has its own unique characteristics: small AAAs might rupture regardless of sac morphology or sex, although this is very rare, while the rupture risk of medium AAAs can be greatly affected by saccular morphology rather than sac diameter, and the rupture risk of large AAAs can be affected by sac diameter rather than sac morphology. Regarding small AAAs, surveillance at intervals of several years is clinically acceptable for men with AAAs in the range of 30 to 40 mm.^19^ Thus, conservative management is generally recommended for patients with small AAAs.^20^ Consistent with this approach, our data showed that there was a small number of non-ruptured fusiform AAAs with diameters < 40 mm, hence the numbers of fusiform and saccular non-ruptured AAAs in these categories were similar. Therefore, the rupture rate in small fusiform AAAs must have been much lower. On the other hand, large AAAs are uncontroversially indicated for repair. Perhaps the most controversial category is medium AAAs, particularly when taking into consideration the contribution of sac morphology to the risk of rupture. At least, since the percentage of saccular AAAs in the 45-49 mm and 50-54 mm categories in this study (6.3% and 3.8%, respectively) was similar to the percentage that was previously reported in AAAs of all sizes, we believe that the statistical analysis for medium AAAs is reasonable.

ROC analysis of our data indicates that aneurysm diameter has an acceptable predictive power for assessing the risk of rupture in both fusiform and saccular AAAs, but it was better in fusiform AAAs. This finding suggests that aneurysm diameter may contribute less to the risk of rupture in saccular AAAs. When considering the size threshold for intervention in AAAs, sensitivity is more important than predictive power itself, because we must avoid false negatives, that is, unexpected rupture. Based on our data, the sensitivity of a cut-off diameter of 55 mm in fusiform AAAs, which is widely accepted as an indication for repair and is reasonable to adopt as a historical control, was comparable with that of a cut-off diameter of 43 mm in saccular AAAs, i.e., 12 mm smaller in saccular AAAs than in fusiform AAAs. Furthermore, the cut-off diameter to predict rupture was 9.0 mm smaller in saccular AAAs than in fusiform AAAs. Therefore, we suggest that the threshold diameter for intervention of saccular AAAs can be set 1 cm smaller than that of fusiform AAAs, although it goes without saying that a size threshold cannot be definitively determined based solely on the findings of the present study.

From a biomechanical perspective, the role of aneurysm geometry in rupture potential has been investigated in the last two decades. The results of previous reports on the effect of aneurysm geometry on mechanical wall stress using finite element analysis were controversial.^6, 21, 22^ Subsequently, using computational fluid dynamics analysis, Boyd et al. reported that aneurysms tended to rupture at the site of low wall shear stress (WSS),^23^ and Natsume et al. proposed that saccular aneurysms with sac depth/neck width > 0.8 had low WSS regardless of diameter, while in fusiform aneurysms WSS was lower as diameter increased.^24^ This may be reflected in our finding that aneurysm diameter may contribute less to the risk of rupture than aneurysm morphology in medium AAAs, and also our finding that the proportion of women was similar between ruptured and non-ruptured cases in saccular AAAs. Akai et al. attempted to identify the subgroup of saccular aneurysms which were truly at high risk of rupture and then defined horizontally long aortic aneurysms with an aspect ratio (neck width/horizontal diameter) < 1.0 as true “saccular” aneurysms.^25^ This study was conducted on thoracic aortic aneurysms, followed by a study on AAAs which revealed that ruptured AAAs had a horizontally longer shape with a smaller fillet radius than non-ruptured AAAs.^26^ In addition, aneurysm shape in the NCD was confirmed by each vascular surgeon, and their judgements were subjective and lacked a detailed definition, except for focal or asymmetric enlargement of the aorta. Hanada et al. reported that a discrepancy existed between a vascular surgeon’s subjective diagnosis and an objective diagnosis using a mechanical structural analysis for AAAs.^27^ As mentioned above, the present study revealed that the diameter of fusiform AAAs has more predictive power for rupture than the diameter of saccular AAAs. We believe that further research using biomechanical approaches will provide a more detailed understanding of the rupture potential of the saccular morphology.

The present study had several limitations. This is not a prospective study that followed preoperative AAA patients from the time when their AAA diameters were small. Therefore, all AAA patients being managed by surveillance or who died before arriving at the operating room have been excluded. In addition, our study only focused on EVAR cases because the NCD did not capture AAA morphology in patients who underwent OSR. Thus, the natural history of saccular AAAs is still unclear. The rupture rate described in our study was the proportion of rupture cases to all EVAR cases performed for AAAs, and this cannot be extrapolated to the true rupture rate of AAAs, especially those with smaller diameters. Growth rate is commonly considered to be a risk for rupture, but was not captured in the NCD, and therefore was not included in the multivariable analysis. There were no specific criteria for the diagnosis of saccular configuration of AAA, so saccular AAAs in our data could be morphologically heterogenous as mentioned above. This study focused on degenerative AAA, but the etiology of saccular AAAs is sometimes difficult to discern. Finally, although the overall sample size was quite large, rupture cases were very rare in small diameter categories and the statistical analysis might not have had sufficient power.

## CONCLUSIONS

Saccular AAAs are more prone to rupture than fusiform AAA in the 40-54 mm diameter range, which supports the current idea that saccular AAAs should be treated at smaller diameters. The 9.0 mm difference in the predicted diameters for the rupture between fusiform and saccular AAAs suggests that the threshold diameter for intervention of saccular AAAs can be set approximately 1 cm smaller than that of fusiform AAAs.

## Data Availability

According to NCD's (NCD; national clinical database) data policy, individual data are provided only to designated analysts.

## ACKNOWLEDGEMENTS

The study was supported by the Japanese Society of Vascular Surgery (JSVS). The authors would like to express their gratitude to Dr. Kimihiro Komori (Chief Director of the JSVS), Dr. Yoshikatsu Saiki (Chief of the Clinical Research Promotion Committee of the JSVS), Dr. Hideaki Obara (Chief of the Database Management Committee of the JSVS), and the staff of the NCD. We would also like to thank all the hospitals participating in this NCD project for their continued efforts concerning data entry.

---

Table 4. Cut-off values, sensitivity and specificity of aneurysm diameter for predicting rupture in fusiform and saccular abdominal aortic aneurysms.

